# SARS-CoV-2 vaccine effectiveness in immunosuppressed kidney transplant recipients

**DOI:** 10.1101/2021.08.07.21261578

**Authors:** Hiam Chemaitelly, Sawsan AlMukdad, Jobin Paravila Joy, Houssein H. Ayoub, Hadi M. Yassine, Fatiha M. Benslimane, Hebah A. Al Khatib, Patrick Tang, Mohammad R. Hasan, Peter Coyle, Zaina Al Kanaani, Einas Al Kuwari, Andrew Jeremijenko, Anvar Hassan Kaleeckal, Ali Nizar Latif, Riyazuddin Mohammad Shaik, Hanan F. Abdul Rahim, Gheyath K. Nasrallah, Mohamed Ghaith Al Kuwari, Adeel A. Butt, Hamad Eid Al Romaihi, Mohamed H. Al-Thani, Mohamad M. Alkadi, Omar Ali, Muna Al-Maslamani, Roberto Bertollini, Hassan Al Malki, Yousuf Almaslamani, Laith J. Abu-Raddad, Abdullatif Al Khal

## Abstract

COVID-19 vaccine protection against infection in immunosuppressed solid organ transplant recipients is unknown but possibly weak with the low proportion of these patients mounting a robust humoral and cellular immune response after vaccination. Using a retrospective cohort study design with cross-over, we assessed vaccine effectiveness among 782 kidney transplant recipients registered at Hamad Medical Corporation, the national public healthcare provider in Qatar, where the BNT162b2 (Pfizer-BioNTech) and mRNA-1273 (Moderna) vaccines have been used in the national immunization campaign. Vaccine effectiveness against any SARS-CoV-2 infection was estimated at 46.6% (95% CI: 0.0-73.7%) ≥14 days after the second dose, 66.0% (95% CI: 21.3-85.3%) ≥42 days after the second dose, and 73.9% (95% CI: 33.0-89.9%) ≥56 days after the second dose. Vaccine effectiveness against any severe, critical, or fatal COVID-19 disease was estimated at 72.3% (95% CI: 0.0-90.9%) ≥14 days after the second dose, 85.0% (95% CI: 35.7-96.5%) ≥42 days after the second dose, and 83.8% (95% CI: 31.3-96.2%) ≥56 days after the second dose. Most vaccine breakthrough infections occurred in the first few weeks after receiving the first and/or second dose. Vaccine effectiveness reached considerable levels in kidney transplant recipients, but vaccine protection mounted slowly and did not reach a high level until several weeks after the second dose.

## Introduction

Coronavirus Disease 2019 (COVID-19) vaccine protection against infection in the immunosuppressed solid organ transplant recipients is unknown but suspected to be weak.^1,2^ Evidence indicates low proportion of these patients mounting a robust humoral and cellular immune response after vaccination.^3-18^ In this investigation, we assessed vaccine effectiveness in immunosuppressed kidney transplant recipients in Qatar, where the BNT162b2^19^ (Pfizer-BioNTech) and mRNA-1273^20^ (Moderna) vaccines have been the vaccines of choice in the national immunization campaign.^21-24^

## Methods

### Study population and databases

We analyzed the kidney transplant database and the national federated Qatar severe acute respiratory syndrome coronavirus 2 (SARS-CoV-2) databases compiled at Hamad Medical Corporation, the main public healthcare provider and the nationally designated provider for all Coronavirus Disease 2019 (COVID-19) healthcare needs. Data were extracted directly from the integrated nationwide digital-health information platform that has captured all SARS-CoV-2-related data in the population of Qatar since the start of the epidemic, including demographic characteristics, all records of polymerase-chain-reaction (PCR) testing (Supplementary Text 1), antibody testing, vaccination, COVID-19 hospitalizations, and classification of infection severity and deaths per World Health Organization (WHO) guidelines (performed by trained medical personnel through individual chart reviews)^25,26^ (Supplementary Text 2). These databases are complete, with no missing information, since every test, vaccination, and hospitalization for SARS-CoV-2 in Qatar is registered using the Hamad Medical Corporation Number (national and universal healthcare system number) and Qatar Identification Number in the national centralized SARS-CoV-2 databases.

### Study design

We performed a retrospective cohort study with a cross-over design to estimate, for kidney transplant recipients, effectiveness of vaccination against SARS-CoV-2 infection as a primary outcome. The study compared incidence of infection in vaccinated persons and those unvaccinated between February 1-July 21, 2021, at a time when the country was undergoing two back-to-back SARS-CoV-2 waves dominated by the B.1.1.7 (Alpha^27^) and B.1.351 (Beta^27^) variants.^21,23,28-30^ It is noteworthy that the first SARS-CoV-2 epidemic wave in Qatar occurred (before introduction of any variant of concern) and peaked in late May, 2020.^31,32^ The second wave was triggered by introduction and expansion of the B.1.1.7 variant and peaked in early March, 2021.^21,23,28,29^ The third wave was dominated by the B.1.351 variant, and peaked in the first week of April, 2021.^21,23,28,29^ The B.1.617.2 (Delta^27^) variant has been introduced more recently in Qatar, and as of July 28, 2021, it remains at low incidence.^28,29^ There is no evidence that any other variant of concern is or has been responsible for appreciable community transmission in Qatar.^28,29^

Individuals in the cohort of kidney transplant recipients were eligible for inclusion in the analysis if they had no prior PCR-confirmed diagnosis and were alive at the start of the follow-up time. The time of follow up was from February 1 up to July 21, 2021. Individuals were followed from February 1, 2021 until documented (PCR-confirmed) infection, death, vaccination (for those unvaccinated), or end-of-study censoring (set at July 21, 2021). Reporting of the study followed the STROBE guidelines (Supplementary Table 1).

### Statistical analysis

Descriptive statistical analysis (frequencies and measures of central tendency) was performed to describe vaccinated and unvaccinated cohorts. Covariate differences between cohorts were evaluated using t-tests for continuous variables and chi-square tests for categorical variables, with two-sided p-value<0.05 indicating a statistically significant association.

Infection hazard ratios and associated 95% confidence interval (CIs) were estimated using regressions applying the Fine-Gray model,^33^ with adjustment for age, sex, and nationality group, to control for differences in exposure risk^31,32,34-37^ and variant exposure^21,28,29^ by sex, age, and nationality. This was done using the STATA 17.0^38^ *stcrreg* command to account further for potential bias arising from the competing risks of vaccination and death,^33^ as the duration of follow up coincided with the rapid scale-up of vaccination in Qatar. Hazard ratios were subsequently used to derive vaccine effectiveness and its 95% CI using the equation, 1-hazard ratio. The 95% CIs were not adjusted for multiplicity. Interactions were not considered.

Adjusted curves for cumulative incidence were generated using the *stcurve* command, which fits a Fine-Gray model,^33^ with adjustment for age, sex, and nationality group, to account for potential bias arising from the competing risks of vaccination and death. In addition to the baseline analysis comparing incidence of documented infection in individuals who completed ≥14 days after the second vaccine dose to that in the cohort of individuals with no vaccination record, two subgroup analyses were performed contrasting incidence in individuals who completed ≥42 days and ≥56 days after the second vaccine dose with that in the unvaccinated cohort.

In all analyses, nationality was categorized into: Bangladeshis, Indians, Nepalese, Pakistanis, and Sri Lankans (Group 1), Qataris (Group 2), and all other nationalities (Group 3). This categorization was informed by the observed epidemiology and exposure risk assessments of SARS-CoV-2 infection in Qatar.^31,32,34-37^ Of note that Qatar has unique demographics where expatriates from over 150 countries comprise 89% of the population.^31,39^

Incidence rate of documented infection was calculated by dividing the number of infection cases identified during the study by the number of person-weeks contributed by all eligible individuals in the vaccinated and unvaccinated cohorts. The incidence rate and associated 95% CI were estimated using a Poisson log-likelihood regression model with the *stptime* command. Follow-up person-time was calculated from study onset (February 1, 2021) until PCR-confirmed diagnosis of infection, all-cause death, vaccination (for those unvaccinated), or end-of-study censoring (July 21, 2021). Statistical analyses were conducted in STATA/SE version 17.0.^38^

### Ethical approval

The study was approved by the Hamad Medical Corporation and Weill Cornell Medicine-Qatar Institutional Review Boards with waiver of informed consent.

## Results

### Study population

Incidence of documented SARS-CoV-2 infection, symptomatic or asymptomatic, was assessed in all immunosuppressed kidney transplant recipients (n=782) registered and clinically followed at Hamad Medical Corporation, the national public healthcare provider in Qatar (Figure 1).

**Figure 1.**
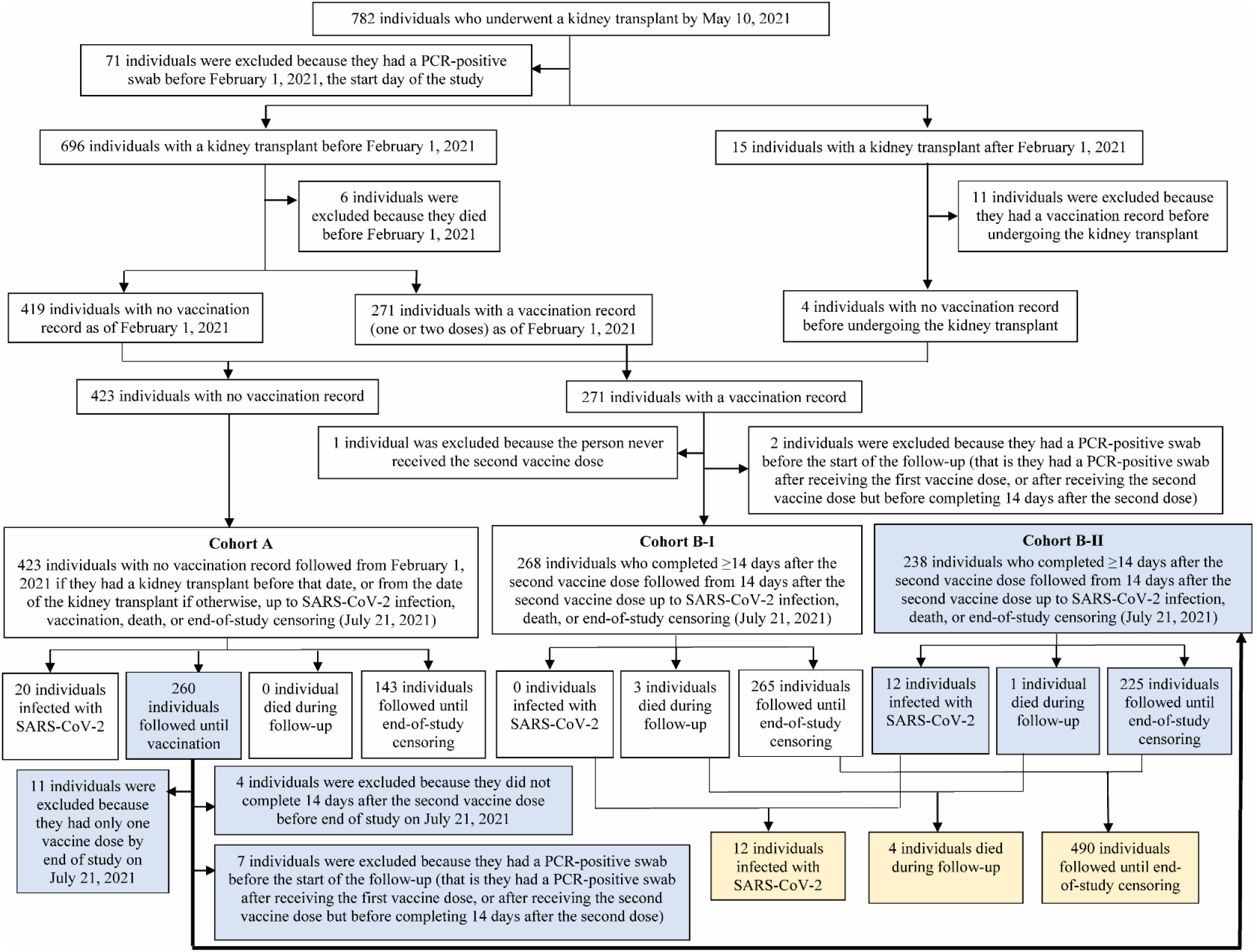
Flowchart showing the selection process for the study population. Light blue color indicates persons who entered the study unvaccinated, but who were subsequently vaccinated and followed up as part of the cohort of vaccinated individuals. Light yellow color indicates final outcomes for the full Cohort B of vaccinated individuals, including Cohort B-I and Cohort B-II.

Maintenance immune suppression regimens included azathioprine (9.6%), cyclosporine (16.4%), everolimus (0.2%), mycophenolate mofetil (76.1%), prednisolone (82.5%), sirolimus (4.2%), and tacrolimus (72.9%). Out of the 782 transplant recipients, 601 (76.9%) received at least one vaccine dose by the end of the study on July 21, 2021, of whom 559 (93.0%) were vaccinated with BNT162b2 and 42 (7.0%) with mRNA-1273. The median date of first vaccine dose was February 2, 2021; and 48.3% of individuals received one or both vaccine doses before start of follow-up on February 1, 2021.

Individuals in this cohort were followed from February 1, 2021 until documented (PCR-confirmed) infection, all-cause death, vaccination (for those unvaccinated), or end-of-study censoring (set at July 21, 2021; Figure 1). Individuals with a prior SARS-CoV-2 PCR-confirmed diagnosis (n=71), or those who died before start of follow-up (n=6), were excluded from analysis.

Figure 1 shows the process for identifying infections in unvaccinated and vaccinated persons, and Table 1 presents their demographic and clinical characteristics. The median time since transplantation was 7 years (interquartile range (IQR): 4-13). The median age of those unvaccinated was 49 years (IQR: 39-61) and 63.1% were men, whereas among those vaccinated, the median age was 52 years (IQR: 40-61) and 70.4% were men. Unvaccinated and vaccinated persons came from diverse national origins and were broadly representative of the unique demographics of the population of Qatar.^31,39^

**Table 1.** Demographic and clinical characteristics of study cohorts.

| Characteristics | Individuals with no vaccination record (Cohort A) | Individuals who completed $\geq 14$ days after the second vaccine dose (Cohort B)* | p-value |
| --- | --- | --- | --- |
| **Median age (IQR) — years** | 49 (39-61) | 52 (40-61) | 0.174 |
| **Age group — no. (%)** |  |  |  |
| <30 years | 34 (8.0) | 39 (7.7) | 0.282 |
| 30-39 years | 76 (18.0) | 74 (14.6) |  |
| 40-49 years | 104 (24.6) | 105 (20.8) |  |
| 50-59 years | 89 (21.0) | 133 (26.3) |  |
| 60-69 years | 88 (20.8) | 116 (22.9) |  |
| 70+ years | 32 (7.6) | 39 (7.7) |  |
| **Sex** |  |  |  |
| Men | 267 (63.1) | 356 (70.4) | 0.019 |
| Women | 156 (36.9) | 150 (29.6) |  |
| **Nationality†** |  |  |  |
| Bangladeshi | 11 (2.6) | 8 (1.6) | 0.005 |
| Egyptian | 31 (7.3) | 30 (5.9) |  |
| Filipino | 27 (6.4) | 30 (5.9) |  |
| Indian | 47 (11.1) | 45 (8.9) |  |
| Nepalese | 2 (0.5) | 3 (0.6) |  |
| Pakistani | 22 (5.2) | 24 (4.7) |  |
| Qatari | 168 (39.7) | 236 (46.6) |  |
| Sri Lankan | 4 (1.0) | 3 (0.6) |  |
| Sudanese | 40 (9.5) | 43 (8.5) |  |
| Other nationalities | 71 (16.8)‡ | 84 (16.6)‡ |  |
| **Median time since transplant (IQR) — years** | 7 (4-13) | 7 (4-13) | 0.261 |
| **Time since transplant** |  |  |  |
| <3 years | 60 (14.2) | 58 (11.5) | 0.651 |
| 3-6 years | 136 (32.2) | 172 (34.0) |  |
| 7-11 years | 98 (23.2) | 121 (23.9) |  |
| $\geq 12$ years | 129 (30.5) | 155 (30.6) | |
| **Maintenance immunosuppression** |  |  |  |
| Azathioprine |  |  |  |
| Yes | 40 (9.5) | 49 (9.7) | 0.907 |
| No | 383 (90.5) | 457 (90.3) |  |
| Cyclosporine |  |  |  |
| Yes | 67 (15.8) | 85 (16.8) | 0.694 |
| No | 356 (84.2) | 421 (83.2) |  |
| Everolimus |  |  |  |
| Yes | 0 (0.0) | 2 (0.4) | 0.196 |
| No | 423 (100.0) | 504 (99.6) |  |
| Mycophenolate mofetil |  |  |  |
| Yes | 303 (71.6) | 404 (79.8) | 0.003 |
| No | 120 (28.4) | 102 (20.2) |  |
| Prednisolone |  |  |  |
| Yes | 333 (78.7) | 433 (85.6) | 0.006 |
| No | 90 (21.3) | 73 (14.4) |  |
| Sirolimus |  |  |  |
| Yes | 19 (4.5) | 20 (4.0) | 0.683 |
| No | 404 (95.5) | 486 (96.1) |  |
| Tacrolimus |  |  |  |
| Yes | 289 (68.3) | 388 (76.7) | 0.004 |
| No | 134 (31.7) | 118 (23.3) |  |

Abbreviations: IQR, interquartile range.
\*This group includes all vaccinated individuals who completed $\geq 14$ days after the second vaccine dose in the cross-over design.
†Nationalities were chosen to represent the most populous groups in the population of Qatar.
‡These comprise 18 other nationalities among individuals with no vaccination record, and 24 other nationalities among individuals who completed $\geq 14$ days after the second vaccine dose.

### Vaccine effectiveness at ≥14 days after the second dose

Figure 2 shows the cumulative incidence of infection among those who completed ≥14 days after the second vaccine dose compared to that in persons with no vaccination record, adjusting for age, sex, nationality group, and the competing risks of vaccination and death. Cumulative incidence in vaccinated and unvaccinated cohorts was assessed, respectively, at 2.58% (95% confidence interval (CI): 1.41-4.35%) versus 4.74% (95% CI: 3.00-7.07%), after 120 days of follow-up. Incidence rate of infection was estimated, respectively, at 13.35 (95% CI: 7.58-23.51) versus 37.73 (95% CI: 24.34-58.48) per 10,000 person-weeks over a total follow-up time of 8,989.1 and 5,301.0 person-weeks, respectively.

**Figure 2.**
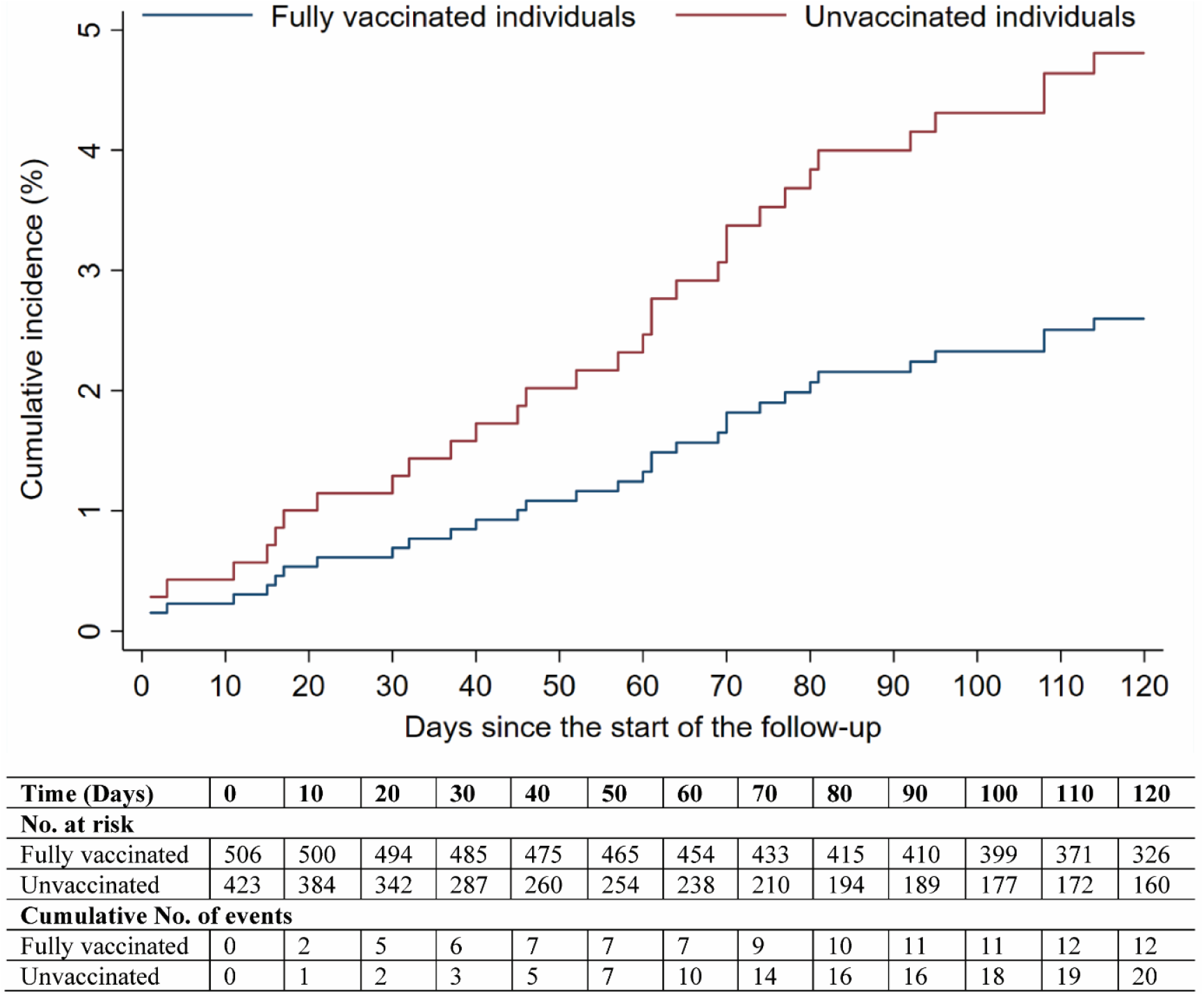
Cumulative risk (incidence) of documented SARS-CoV-2 infection in the cohort of vaccinated individuals who completed ≥14 days after the second vaccine dose, compared to that of documented infection in the cohort of individuals with no vaccination record, calculated using the Fine-Gray model to account for the competing risks of vaccination and death, and after adjustment for age, sex, and nationality group. Follow-up extended from February 1 or the organ transplant date (which ever came first) for the cohort of unvaccinated individuals, and from 14 days after the second vaccine dose for the cohort of vaccinated individuals, both up to July 21, 2021.

The hazard ratio adjusted for age, sex, nationality group, and the competing risks of vaccination and death was 0.53 (95% CI: 0.26-1.08). Vaccine effectiveness against any SARS-CoV-2 infection, symptomatic or asymptomatic, was derived using the equation, 1-hazard ratio, and was estimated at 46.6% (95% CI: 0.0-73.7%). Vaccine effectiveness against any severe, critical, or fatal COVID-19 disease was estimated at 72.3% (95% CI: 0.0-90.9%).

### Vaccine effectiveness at ≥42 days and ≥56 days after the second dose

Analyses were repeated to estimate vaccine effectiveness at ≥42 days and at ≥56 days after the second dose, instead of at ≥14 days after the second dose. Figures 3 and 4 show the cumulative incidence of infection including only the cohorts of individuals who completed ≥42 days and ≥56 days after the second vaccine dose, respectively, compared to that in the cohort of individuals with no vaccination record, adjusting for age, sex, nationality group, and the competing risks of vaccination and death. Cumulative incidence was assessed, respectively, at 1.60% (95% CI: 0.72-3.15%) and 1.19% (95% CI: 0.45-2.63%) among those vaccinated, versus 4.74% (95% CI: 3.00-7.07%) among those unvaccinated, after 120 days of follow-up.

**Figure 3.**
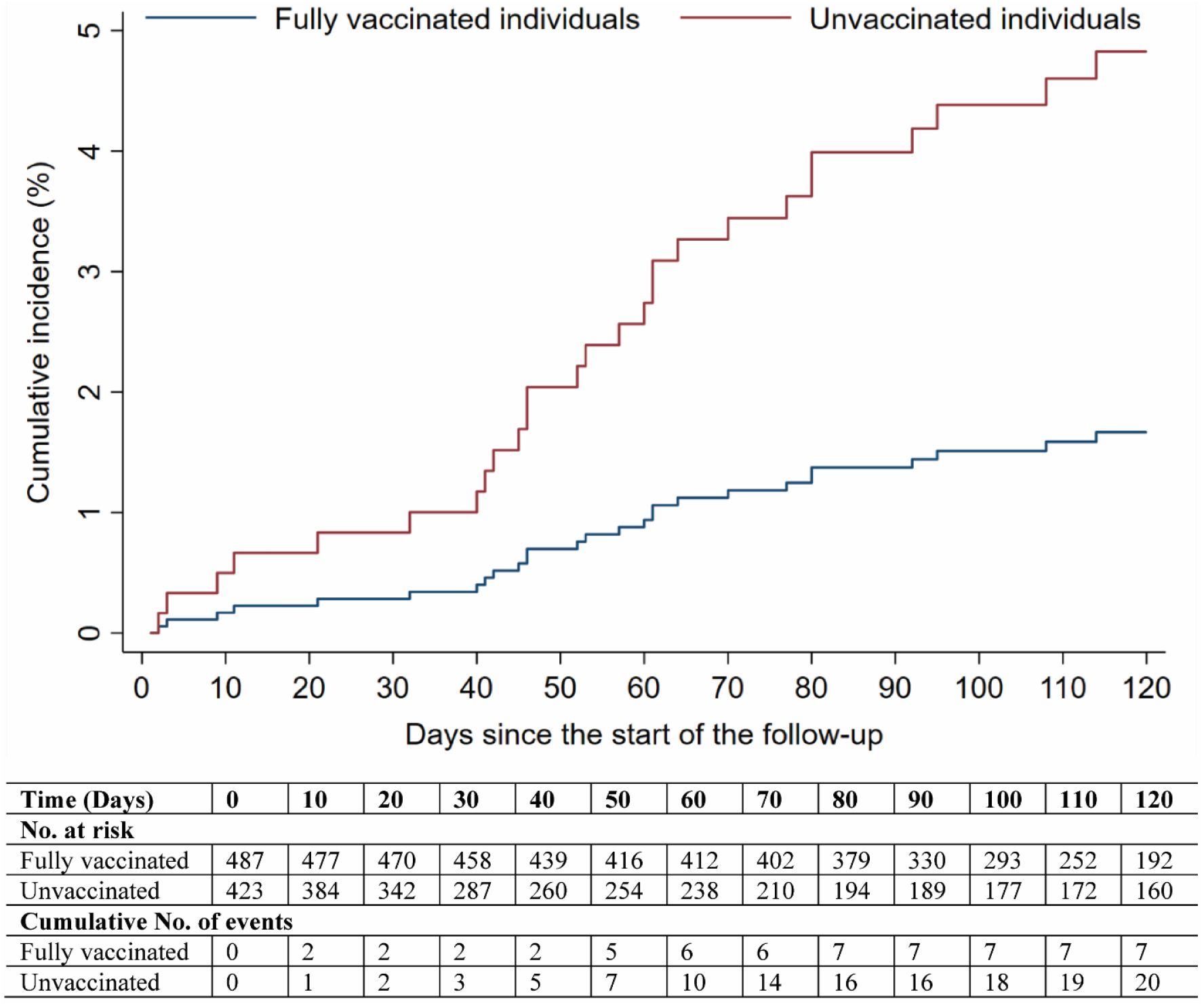
Cumulative risk (incidence) of documented SARS-CoV-2 infection in the cohort of vaccinated individuals who completed ≥42 days after the second vaccine dose, compared to that of documented infection in the cohort of individuals with no vaccination record, calculated using the Fine-Gray model to account for the competing risks of vaccination and death, and after adjustment for age, sex, and nationality group. Follow-up extended from February 1 or the organ transplant date (which ever came first) for the cohort of unvaccinated individuals, and from 42 days after the second vaccine dose for the cohort of vaccinated individuals, both up to July 21, 2021.

**Figure 4.**
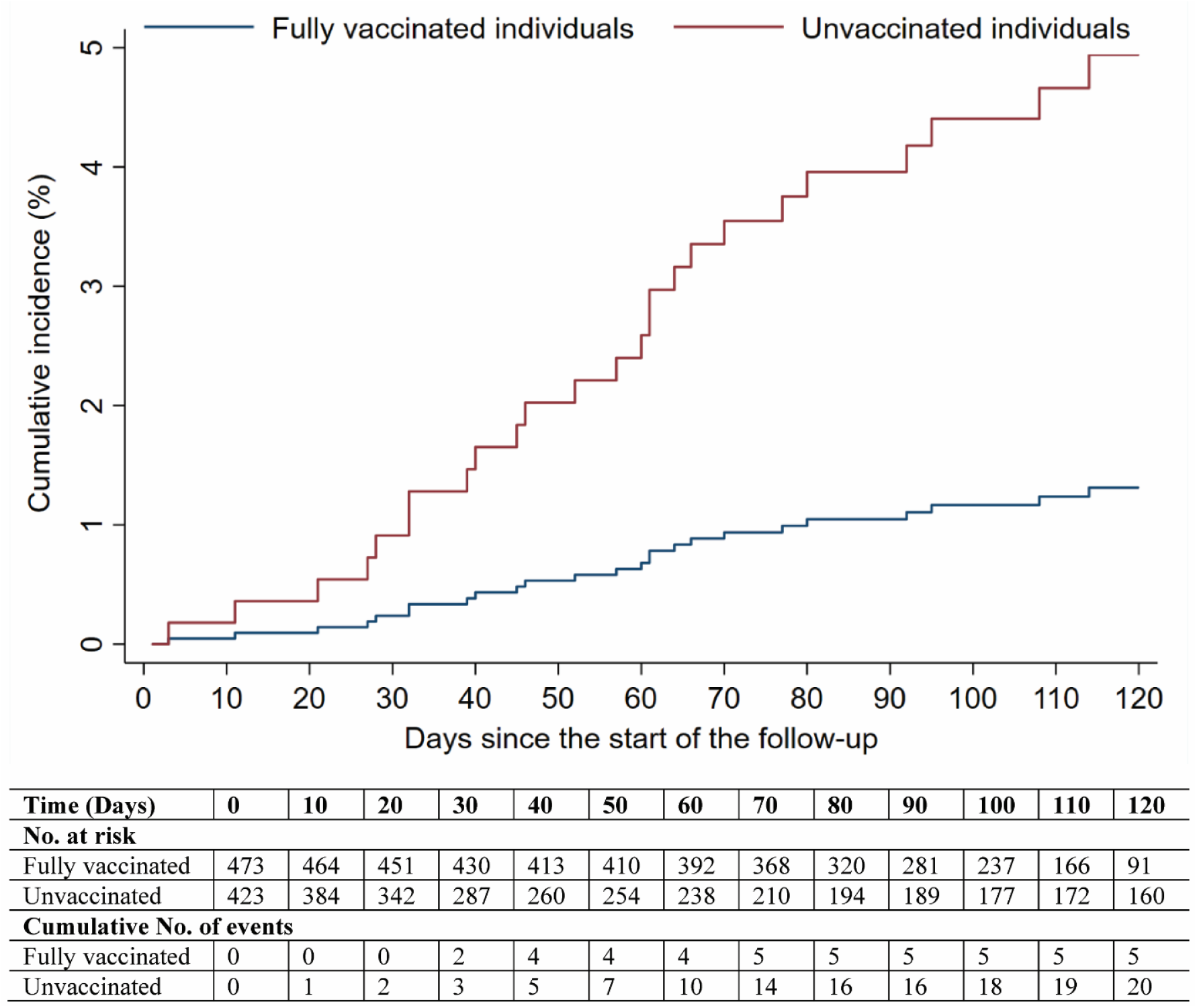
Cumulative risk (incidence) of documented SARS-CoV-2 infection in the cohort of vaccinated individuals who completed ≥56 days after the second vaccine dose, compared to that of documented infection in the cohort of individuals with no vaccination record, calculated using the Fine-Gray model to account for the competing risks of vaccination and death, and after adjustment for age, sex, and nationality group. Follow-up extended from February 1 or the organ transplant date (which ever came first) for the cohort of unvaccinated individuals, and from 56 days after the second vaccine dose for the cohort of vaccinated individuals, both up to July 21, 2021.

Incidence rates of infection in the cohorts of individuals who completed ≥42 days and ≥56 days after the second vaccine dose and the cohort of individuals with no vaccination record, were estimated, respectively, at 9.99 (95% CI: 4.76-20.95) and 8.26 (95% CI: 3.44-19.86) per 10,000 person-weeks over a total follow-up time of 7,007.7 and 6,049.9 person-weeks, respectively, among those vaccinated, versus 37.73 (95% CI: 24.34-58.48) per 10,000 person-weeks over a total follow-up time of 5,301.0 person-weeks among those unvaccinated.

Hazard ratios adjusted for age, sex, nationality group, and the competing risks of vaccination and death were estimated at 0.34 (95% CI: 0.15-0.79) and 0.26 (95% CI: 0.10-0.67), respectively. Vaccine effectiveness against any SARS-CoV-2 infection, symptomatic or asymptomatic, was estimated at 66.0% (95% CI: 21.3-85.3%) in those who completed ≥42 days, and at 73.9% (95% CI: 33.0-89.9%) in those who completed ≥56 days. Vaccine effectiveness against any severe, critical, or fatal COVID-19 disease was estimated at 85.0% (95% CI: 35.7-96.5%) in those who completed ≥42 days, and at 83.8% (95% CI: 31.3-96.2%) in those who completed ≥56 days.

## Discussion

Vaccine effectiveness reached considerable levels in immunosuppressed kidney transplant recipients, which were not far below those in the general population of Qatar.^21-24^ However, vaccine protection mounted slowly and did not reach a high level until several weeks after the second dose. Notably, the build-up of vaccine protection mirrored the slow development of antibodies in transplant recipients that has been previously reported.^3,5-9,11-14,16^ Indeed, the majority of breakthrough infections in those vaccinated occurred in the first few weeks after receiving the first and/or second dose (note Figure 1 and events of infection among those vaccinated in Figure 2). Strikingly, no breakthrough infection was recorded in those who were already fully vaccinated before February 1, 2021, the start day of follow-up in this study (Cohort B-I in Figure 1).

Vaccine effectiveness appeared stronger against hospitalization and death than against infection, a finding consistent with the pattern seen in the general population.^21,23^ Of 12 breakthrough infections documented among those vaccinated, four progressed to severe disease, none to critical disease, and none to COVID-19 death. Meanwhile, of 20 infections documented among unvaccinated persons, seven progressed to severe disease, six to critical disease, and none to COVID-19 death. Severe disease (acute-care hospitalization), critical disease (ICU hospitalization), and COVID-19 death were defined per World Health Organization guidelines^25,26^ (Supplementary Text 2).

This study has limitations. With the relatively small cohort of transplant recipients (n=782), and the rapid scale-up of vaccination during the study duration, it was not possible to match vaccinated and unvaccinated persons by factors such as age, sex, nationality, comorbidity, and calendar time, while maintaining adequate sample sizes, time of follow-up, and statistical precision. However, cohorts of both vaccinated and unvaccinated were largely balanced by both demographic and clinical characteristics (Table 1). Moreover, the regression analyses adjusted for age, sex, and nationality group, with age being a proxy for comorbidity, and both the adjusted and unadjusted analyses reached similar results (not shown). Additionally, the majority of individuals in both vaccinated and unvaccinated cohorts were followed during similar times, starting from the first day of the study, February 1, 2021, or shortly thereafter, even in the additional analyses of vaccine effectiveness for those at ≥42 days and at ≥56 days after the second vaccine dose. Please note follow up times (results and Figure 1) and numbers at risk (Figures 2-4).

We only assessed risk of documented infection, but other infections may have occurred and gone undocumented, perhaps because of minimal/mild or no symptoms. Unlike in blinded, randomized clinical trials, the investigated observational cohorts were not blinded nor randomized. Vaccine effectiveness estimates calculated in this study may not necessarily be generalizable to other solid organ transplant populations.

In conclusion, despite these limitations, the findings of this study demonstrate build-up of vaccine-induced protection in kidney transplant recipients, against both SARS-CoV-2 infection and COVID-19 disease, but this protection mounts slowly and does not reach a high level until several weeks after the second vaccine dose. While the present findings indicate that even immunosuppressed individuals benefit from vaccination, they suggest the need to develop better strategies to enhance the protection in this group, such as exploring the addition of a third vaccine dose.

## Data Availability

The dataset of this study is a property of the Qatar Ministry of Public Health that was provided to the researchers through a restricted-access agreement that prevents sharing the dataset with a third party or publicly. Future access to this dataset can be considered through a direct application for data access to Her Excellency the Minister of Public Health (https://www.moph.gov.qa/english/Pages/default.aspx). Aggregate data are available within the manuscript and its Supplementary information.

## Acknowledgements

We acknowledge the many dedicated individuals at Hamad Medical Corporation, the Ministry of Public Health, the Primary Health Care Corporation, and the Qatar Biobank for their diligent efforts and contributions to make this study possible. The authors are grateful for support from the Biomedical Research Program, the Biostatistics, Epidemiology, and Biomathematics Research Core, and the Genomics Core, all at Weill Cornell Medicine-Qatar, as well as for support provided by the Ministry of Public Health and Hamad Medical Corporation. The authors are also grateful for support from the Qatar Genome Programme for supporting the viral genome sequencing. Statements made herein are solely the responsibility of the authors. The funders of the study had no role in study design, data collection, data analysis, data interpretation, or writing of the article.

## Author contributions

HC designed the study, performed the statistical analyses, and co-wrote the first draft of the article. LJA co-conceived and co-designed the study, led the statistical analyses, and co-wrote the first draft of the article. AAK co-conceived and co-designed the study. AAK, YA, HAM, OA, MMA, and JPJ constructed and populated the database of kidney transplant recipients. All authors contributed to data collection and acquisition, database development, discussion and interpretation of the results, literature reviews, and to the writing of the manuscript. All authors have read and approved the final manuscript.

## Supplementary Material

### Supplementary Text 1. Laboratory methods

Nasopharyngeal and/or oropharyngeal swabs (Huachenyang Technology, China) were collected for PCR testing and placed in Universal Transport Medium (UTM). Aliquots of UTM were: extracted on a QIAsymphony platform (QIAGEN, USA) and tested with RT-qPCR using TaqPath(tm) COVID-19 Combo Kits (100% sensitivity and specificity;^1^ Thermo Fisher Scientific, USA) on an ABI 7500 FAST (ThermoFisher, USA); extracted using a custom protocol^2^ on a Hamilton Microlab STAR (Hamilton, USA) and tested using AccuPower SARS-CoV-2 Real-Time RT-PCR Kits (100% sensitivity and specificity;^3^ Bioneer, Korea) on an ABI 7500 FAST; or loaded directly into a Roche cobas® 6800 system and assayed with a cobas® SARS-CoV-2 Test (95% sensitivity, 100% specificity;^4^ Roche, Switzerland). The first assay targets the viral S, N, and ORF1ab regions. The second targets the viral RdRp and E-gene regions, and the third targets the ORF1ab and E-gene regions.

All tests were conducted at the HMC Central Laboratory or Sidra Medicine Laboratory, following standardized protocols.

### Supplementary Text 2. COVID-19 severity, criticality, and fatality classification

Severe COVID-19 disease was defined per WHO classification as a SARS-CoV-2 infected person with “oxygen saturation of <90% on room air, and/or respiratory rate of >30 breaths/minute in adults and children >5 years old (or ≥60 breaths/minute in children <2 months old or ≥50 breaths/minute in children 2–11 months old or ≥40 breaths/minute in children 1–5 years old), and/or signs of severe respiratory distress (accessory muscle use and inability to complete full sentences, and, in children, very severe chest wall indrawing, grunting, central cyanosis, or presence of any other general danger signs)”.^5^ Detailed WHO criteria for classifying SARS-CoV-2 infection severity can be found in the WHO technical report.^5^

Critical COVID-19 disease was defined per WHO classification as a SARS-CoV-2 infected person with “acute respiratory distress syndrome, sepsis, septic shock, or other conditions that would normally require the provision of life sustaining therapies such as mechanical ventilation (invasive or non-invasive) or vasopressor therapy”.^5^ Detailed WHO criteria for classifying SARS-CoV-2 infection criticality can be found in the WHO technical report.^5^

COVID-19 death was defined per WHO classification as “a death resulting from a clinically compatible illness, in a probable or confirmed COVID-19 case, unless there is a clear alternative cause of death that cannot be related to COVID-19 disease (e.g. trauma). There should be no period of complete recovery from COVID-19 between illness and death. A death due to COVID-19 may not be attributed to another disease (e.g. cancer) and should be counted independently of preexisting conditions that are suspected of triggering a severe course of COVID-19”. Detailed WHO criteria for classifying COVID-19 death can be found in the WHO technical report.^6^

**Supplementary Table 1.** STROBE checklist for cohort studies.

|  | Item No | Recommendation | Main text page |
| --- | --- | --- | --- |
| Title and abstract | 1 | (a) Indicate the study’s design with a commonly used term in the title or the abstract | 3 |
|  |  | (b) Provide in the abstract an informative and balanced summary of what was done and what was found | 3 |
| Introduction |  |  |  |
| Background/rationale | 2 | Explain the scientific background and rationale for the investigation being reported | 5 |
| Objectives | 3 | State specific objectives, including any prespecified hypotheses | 5 |
| Methods |  |  |  |
| Study design | 4 | Present key elements of study design early in the paper | 5-6 |
| Setting | 5 | Describe the setting, locations, and relevant dates, including periods of recruitment, exposure, follow-up, and data collection | 5-8, & Supplementary Texts 1-2 |
| Participants | 6 | (a) Give the eligibility criteria, and the sources and methods of selection of participants. Describe methods of follow-up | 5-6 |
|  |  | (b) For matched studies, give matching criteria and number of exposed and unexposed | NA |
| Variables | 7 | Clearly define all outcomes, exposures, predictors, potential confounders, and effect modifiers. Give diagnostic criteria, if applicable | 5-8 & Supplementary Text 2 |
| Data sources/ measurement | 8* | For each variable of interest, give sources of data and details of methods of assessment (measurement). Describe comparability of assessment methods if there is more than one group | 5-8 |
| Bias | 9 | Describe any efforts to address potential sources of bias | 7 |
| Study size | 10 | Explain how the study size was arrived at | 6 & Figure 1 |
| Quantitative variables | 11 | Explain how quantitative variables were handled in the analyses. If applicable, describe which groupings were chosen and why | 7 & Table 1 |
| Statistical methods | 12 | (a) Describe all statistical methods, including those used to control for confounding | 7-8 |
|  |  | (b) Describe any methods used to examine subgroups and interactions | 7 |
|  |  | (c) Explain how missing data were addressed | NA, see p. 5 |
|  |  | (d) If applicable, explain how loss to follow-up was addressed | NA, see p. 5 |
|  |  | (e) Describe any sensitivity analyses | 7 |
| Results |  |  |  |
| Participants | 13* | (a) Report numbers of individuals at each stage of study—eg numbers potentially eligible, examined for eligibility, confirmed eligible, included in the study, completing follow-up, and analysed<br>(b) Give reasons for non-participation at each stage<br>(c) Consider use of a flow diagram | 8 & Figure 1 |
| Descriptive data | 14 | (a) Give characteristics of study participants (eg demographic, clinical, social) and information on exposures and potential confounders | 8-9, Figure 1 & Table 1 |
|  |  | (b) Indicate number of participants with missing data for each variable of interest | NA, see p. 5 |
|  |  | (c) Summarise follow-up time (eg, average and total amount) | 9-10 & Figures 1-4 |
| Outcome data | 15 | Report numbers of outcome events or summary measures over time | 8-11 & Figures 1-4 |
| Main results | 16 | (a) Give unadjusted estimates and, if applicable, confounder-adjusted estimates and their precision (eg, 95% confidence interval). Make clear which confounders were adjusted for and why they were included | 8-11 & Figures 2-4 |
|  |  | (b) Report category boundaries when continuous variables were categorized | NA |
|  |  | (c) If relevant, consider translating estimates of relative risk into absolute risk for a meaningful time period | NA |
| Other analyses | 17 | Report other analyses done—eg analyses of subgroups and interactions, and sensitivity analyses | 10-11 & Figures 3-4 |
| Discussion |  |  |  |
| Key results | 18 | Summarise key results with reference to study objectives | 11 |
| Limitations | 19 | Discuss limitations of the study, taking into account sources of potential bias or imprecision. Discuss both direction and magnitude of any potential bias | 12 |
| Interpretation | 20 | Give a cautious overall interpretation of results considering objectives, limitations, multiplicity of analyses, results from similar studies, and other relevant evidence | 12-13 |
| Generalisability | 21 | Discuss the generalisability (external validity) of the study results | 12 |
| Other information |  |  |  |
| Funding | 22 | Give the source of funding and the role of the funders for the present study and, if applicable, for the original study on which the present article is based | 14 |

NA, not applicable

